# The Black community’s perspectives on the Black maternal health crisis

**DOI:** 10.1101/2025.02.19.25322539

**Authors:** Kaytura Felix, Jay Mawuli, Alejandro McGhee, Marjorie Paloma, Keshia Pollack Porter

**Affiliations:** Department of Health Policy and Management, Bloomberg School of Public Health, Johns Hopkins University, Baltimore, Maryland, United States of America; Department of Strategic Portfolios, Robert Wood Johnson Foundation, Princeton, New Jersey, United States of America

## Abstract

Many Black people in the United States have responded to their excessively high risk for maternal mortality and morbidity by moving childbirth from hospital settings to community homes and birth centers. Black perinatal health leaders are increasingly advocating for investments in community strategies such as birth centers and community midwifery. Despite the excess burden of death and disease carried by that community, few studies have sought to investigate community solutions. This qualitative study was conducted between January and October of 2023 to explore how the Black community was responding to the maternal health crisis. It consisted of structured and unstructured conversations with fifty-seven experts, leaders, artists, and practitioners who were deeply knowledgeable about community approaches and an observation of one Black midwife caring for two Black postpartum clients in their homes. Participants perceived that the overemphasis on the negative mortality and morbidity statistics on Black birthing people in the media, without a concomitant attention to the solutions, is harmful to that community and discourages childbearing. They described significant yet often ignored positive contributions of Black women and identified Black community midwifery as an important yet under-resourced strategy. Direct observation of community midwifery care illustrated dominant themes of spirituality, support, and rest. These findings suggest that Black community midwifery is an important model of care that offers protective factors during the times that Black birthing people —postpartum—are most vulnerable to death and disease and warrant further investments and study.

## Introduction

Maternal mortality is a major public health problem worldwide. As of 2020, the global maternal mortality ratio is 223 maternal deaths per 100,000 live births [1]. Although this ratio has trended downward in the past 30 years [1], it far exceeds the United Nations Sustainable Development Goals of less than 70 deaths per 100,000 live births [2]. Strategies to improve maternal health care can help prevent maternal mortality and reduce maternal morbidity.

Community-based childbirth—childbirth in the home or birth centers— is rapidly growing and is seen as one possible way to prevent these deaths. Between 2004 and 2017, community-based childbirth increased by 77%, and between 2016 and 2023, home birth rates increased by 60% [3, 4]. Black communities are embracing these births more than other populations. Data from the United States Center for Disease Control and Prevention (CDC) indicate that between 2020 and 2021 community-based birth increased by 12% for the general United States population, and by 21% for Black Americans [5]. This small but significant shift reflects in part the safety concerns that many families in the United States had about hospitals during the COVID-19 pandemic [6]. For Black families, it also reflects a response to the maternal health crisis that has been unfolding within the American health care and public health systems—one that disproportionately harms Black people and other people of color [7]. Black birthing people’s interest in and demand for community-based childbirth predates the pandemic and continues in the post-pandemic period.

In 2020, the CDC reported that Black women are at least three times more likely to die during pregnancy compared to their white counterparts [8]. Additionally, more than a third of Black women report mistreatment during maternity care [9] and are more likely, than other racial and ethnic groups, to experience cesarean delivery with its associated risks [10]. Films such as Aftershock [11] and Birthing Justice [12] have documented the impact of the American childbirth system on Black families and their response to the crisis.

Globally, community-based childbirth has been demonstrated to positively impact maternal and newborn mortality rates through improved access to care, enhanced health practices, and effective educational interventions, especially in low resource settings such as low-income countries where traditional healthcare systems may be lacking or inaccessible. [13]. Research also suggests the wide range of United States based community-based models of childbirth could improve maternal health outcomes and patients’ experiences particularly for people of color and those with low incomes [14]. Within the Global South in particular, community midwifery remains an ancient, deep-rooted practice. Women in rural South Asian and African communities overwhelmingly prefer traditional midwifery care. They cited trust, fewer experiences of obstetric violence, and greater emotional and physical safety as reasons for seeking out community birthing care [15].

Until the early1900s most Black people, especially those in the southern United States were born at home, with the assistance of a Black lay midwife. As late as the 1940s, Black midwives attended approximately 75% of births in southern states [16]. Prominent physicians, organized physician groups like the American Medical Association, and public health leaders conducted a national disinformation campaign to discredit midwives, advanced legislations such as the Sheppard Towner Act that disenfranchised lay midwives, and made massive investments in physician and nursing workforce, hospital infrastructure and payment models [16, 17]. Midwifery education was handed off to public health nurses, many of whom had little to no experience in attending births. If Black midwives sought to practice within these constraints, they were required to “submit to the supervision” of these nurses [17]. These efforts resulted in the marginalization of Black community midwifery and the near complete transfer of childbirth to the hospital. By the late 1960s, roughly 1% of US births occurred in the community though data collection on Black community birth remained sparse. [18].

Midwives are the primary providers of community-based childbirth. The midwifery model of care results in improved mother and infant outcomes, including lower rates of preterm birth [19], lower rates of low birth weight [20], higher rates of positive postpartum experiences, including improved self-expression during birth, higher postpartum self-esteem, and greater satisfaction with the birthing experience [21]. Today, the number of community births has hovered around 2% for white Americans and 1% for Black Americans, with Black people seeking this option at an increasing rate [22]. Notably among Black Americans, there has been a longstanding and growing interest in midwifery-led community-based childbirth. A survey of Black women in 2016 revealed a strong desire for community-based childbirth, with 12% of those surveyed noting that they “felt safest giving birth outside the hospital” [22]. However, only a very small proportion of Black women have access to this effective model of care. Barriers to community birth for Black birthing people are numerous and multifaceted, often spanning the domains of financial and legal constraints and lack of community and medical institutional support. Few studies, however, have explored Black community-based birthing. This gap is especially glaring given the excess burden of disease and death that is carried by the Black community and the promise of this effective model of care. We know little about the infrastructure that supports it or the experience of those involved. This article documents early efforts to address this important gap in knowledge on Black community-based childbirth.

## Methods & Approaches

### Participants and Data Collection

This study was conducted, in three phases, as part of a landscape analysis to better understand what actions the Black community was taking to address its maternal health crisis. This study employed an implicit asset frame in that it focused on solutions within the affected community. All participants were informed that they were participating in a conversation that would inform the development of a research study that would contribute to what the Black community was already doing and an advisory group that would guide the research. The study combined purposive and snowball sampling to engage participants from January 23 until October 30, 2023. In the first phase, participants were identified through conversations with peers and articles on Black maternal health in the academic and lay press. Participants were deemed to be deeply knowledgeable about community approaches to the maternal health crisis because they were addressing it directly or supporting those who were addressing it. At the end of each conversation, participants were asked to identify someone who would be knowledgeable about or interested in the topic.

First phase interviews were conducted by the lead author as unstructured conversations and were guided by the following open-ended question: 1) “What was the Black community doing to address the high rates of maternal mortality and morbidity?” Although the interviewer did not ask participants for their demographic information, she noted that information whenever it was mentioned. The conversations were conducted over Zoom video, a cloud-based videoconferencing service [23] and lasted approximately thirty minutes. To establish rapport, none of the conversations were recorded. The lead author took notes during and after the meeting.

The second phase included semi-structured interviews of key figures who emerged from the first phase via snowball sampling. A key figure was someone who was mentioned by two or more people. Because community-based childbirth arose as an important community strategy, this phase sought further exploration of this topic (Table 1). Interviews ranged from thirty-five to forty-five minutes, were held on Zoom video, and were audio-recorded. Participants from small organizations were offered an honorarium.

**Table 1.** Second Phase Interview Questions.

| Questions |
| --- |
| • How are Black communities addressing the high Black maternal morbidity and mortality?<br>• What birthing alternatives/innovations are people in the Black community advancing?<br>• Who are the key players in this space nationally?<br>• Which stakeholders' buy-in are important in the development of community-based births?<br>• What conditions facilitate/hinder the development of community-based birthing?<br>• What is the Black midwife's or birth center's role in addressing the Black maternal health crisis?<br>• What is your experience collaborating with midwives and/or birth centers?<br>• In your experience, which healthcare systems are addressing the Black maternal health crisis? How do they work with communities to address this challenge? |

The third phase included observations of a Black community midwife who was providing care to Black clients. This was deemed necessary because the work of Black community midwives had emerged as an important aspect of the Black community’s response to the Black maternal health crisis, and the team had no prior knowledge of midwifery. The team approached three Black community midwives who cared for Black clients in a region of the country where there were several Black community midwives. One consented to being observed ‘to help [the team] design a study of Black midwives caring for Black clients. The lead author shadowed that midwife on two postpartum home visits on two different days for a total of eight hours. The first was a ten-day postpartum visit and the second was an eight-week visit; both were between ninety minutes and two hours. The midwife obtained the clients’ consent for the observation prior to the visit and reaffirmed the consent at the outset of the visit. Each engagement with the midwife also included time commuting to and from the home visits. Immediately after the visit, the lead author clarified with the midwife sections of the visit that were unclear and then developed a detailed written report of her observations. The midwife received an honorarium.

### Data analysis

The audio recordings were transcribed and checked for accuracy using Otter.ai, a web-based transcription service that utilizes machine learning [24]. The research team reviewed and discussed iteratively hand notes and transcripts to generate inductive codes that were compiled into a simple codebook. Inductive codes were used because of the open-ended nature of the interviews. Two coauthors/coders used Quirkos, a qualitative data analysis software [25], to code the transcripts and identify core themes. The team also reviewed and discussed the observation reports to identify themes.

### Ethics Statement

Human subjects research approval was obtained from Johns Hopkins Bloomberg School of Public Health’s Institutional Review Board (IRB #30521). Informed oral consent was obtained at the outset of the interview and observations.

## Results

Fifty-seven individuals participated in the study between January and October of 2023. (Table 2) The overwhelming majority presented as female and Black; four identified themselves as people of color and four presented or self-identified as White. Participants who voluntarily identified themselves as Black included African Americans, Africans, Afro-Indigenous, Afro-Caribbean, and Afro-Latinas. Participants included home birth and birth center midwives; doulas, an obstetrician, and a maternal-fetal medicine specialist, a lactation specialist; public health researchers; community advocates and activists; and artists.

**Table 2.**
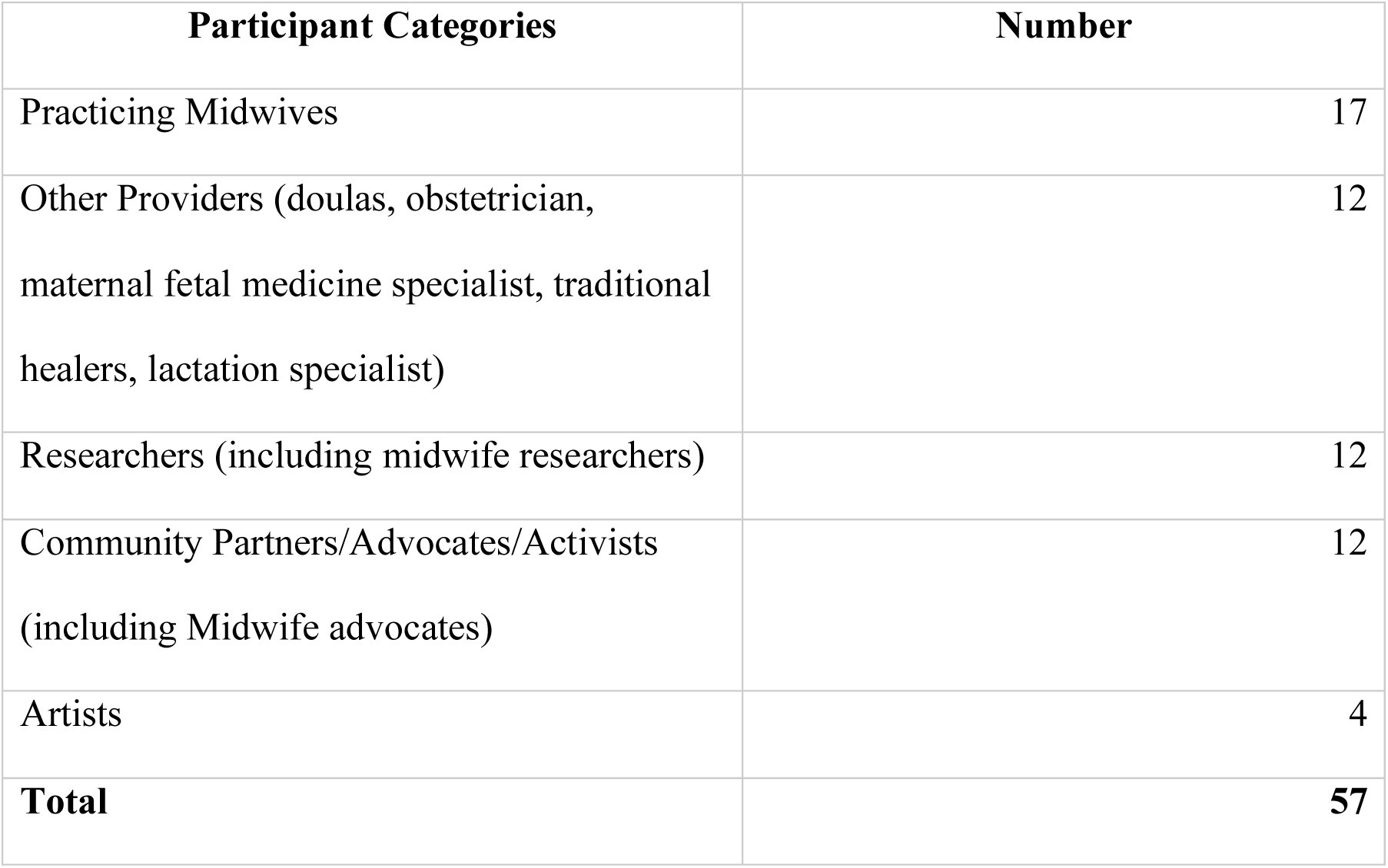
Participant Categories.

| Participant Categories | Number |
| --- | --- |
| Practicing Midwives | 17 |
| Other Providers (doulas, obstetrician, maternal fetal medicine specialist, traditional healers, lactation specialist) | 12 |
| Researchers (including midwife researchers) | 12 |
| Community Partners/Advocates/Activists (including Midwife advocates) | 12 |
| Artists | 4 |
| **Total** | **57** |

Forty-seven and ten individuals participated in the first and second phase respectively. Seven did not respond to our invitation to participate. Most individuals expressed appreciation or enthusiasm for the positive framing of the research and its focus on community assets and solutions to address poor maternal outcomes. Participants offered unprompted statements of appreciation, best wishes for the research, and a desire to stay connected. The results of the interviews and observations are presented below in terms of the major themes that emerged.

### Interviews

#### The overemphasis on negative mortality and morbidity statistics is harmful

Participants expressed frustration about the current negative discourse on Black maternal health in the US. They described a national narrative that was focused primarily on negative statistics, did not look “beyond the numbers,” and overlooked “Black joy” and resourcefulness. Some concluded that there seemed to be more “admiration for the problem” than engaging in solutions.

Respondents went on to argue that the narratives that focus on the negative statistics discouraged pregnancy and birthing altogether in the Black community. These narratives are “[s]caring people…and retraumatizing people…[and] people are scared to have children.” Some also noted that “[f]ear feeds into worse [perinatal] outcomes.”

Participants also suggested that a solution-oriented frame is needed to counterbalance traumatizing statistics and heavy and incomplete narratives about Black maternal health. They cautioned however that solutions must speak to the current crisis and “avoid creat[ing] more trauma.” Regarding the solutions, participants pointed to community assets in rural and immigrant communities that kept the [practices, standards and sanctity of birth.]

#### Centering the voices and investments of the traditionally marginalized frontline workers and leaders—Black women—is just

Individuals emphasized that Black women were working as community leaders to address the maternal health crisis. They also noted that these women’s efforts extended beyond birth and pregnancy into broader justice and equity issues. One individual captured the leadership of Black women, noting: “I think the Black [women in] communities are mobilizing in grassroots efforts to address the maternal health crisis that we’re seeing in our community.” Another described these women’s approach as holistic, noting that “[N]one of their work is just about birth or pregnancy…” Participants pointed out that Black women leaders embraced diverse roles such as advocacy and workforce development, such as training community birth workers to address these challenges.

Participants described a vexing tendency to lionize the Black women leaders on the frontlines of the crisis without an accompanying commitment to supporting them. Acknowledging the voices of Black women as frontline workers and leaders is crucial for addressing the maternal health crisis in Black communities and ensuring birthing people receive the highest quality of care.

#### Black community midwifery is an important yet under-resourced strategy for addressing the crisis

Participants described the care that Black community midwives provided to Black clients as an important strategy for redressing the Black maternal health crisis. Their provision of community-based childbirth care helps the clients avoid the trauma of hospital births. They play an under-appreciated yet crucial role in reshaping maternal health care by offering culturally concordant and meaningful alternatives to the current dominant model of childbirth. As primary providers of community childbirth care, they present opportunities for transformation of the experience that Black people in the U.S have with childbirth. A few expressed frustrations that Black midwives, the practitioners who make community-birthing possible, were generally overlooked in policy conversations at the state and national levels.

Some pointed out that the investments in doulas, while important and necessary, were insufficient to make significant changes because doulas work, within the constraints of the dominant obstetrics model, to mitigate the harm and cannot transform it. Moreover, doulas are themselves victims of the direct or vicarious trauma they experience in hospitals. A consequence of this underinvestment is “[B]lack women of birthing age are [generally] not aware of how meaningful that [community birth] alternatives can be.”

Participants also suggested that the lack of support for Black community midwives who serve their communities places them under stress. The healthcare system is traumatizing Black childbearing people, midwives, and other birth workers. Assessing the current state of Black community midwifery is urgent because “not only are Black patients suffering, but community [midwifery] practitioners are [also] suffering under a broken system that prioritizes capitalism over holistic and integrated healthcare practices.” Another individual noted: “I would like to see all of the professions respected and integrated into one space, and I would like us to be able to practice to our full scope of practice without the restrictive licensure that we have.”

#### A contextual approach to the research is needed

A few individuals noted passionately that community childbirth was occurring ‘within an embedding context of history, society, and policy.” Different actors within the community— midwives and their collaborators—made community-based childbirth possible. Some community organizations ameliorated the harm faced by birthing people. Participants recommended that research be more holistic by taking “a 360-degree look around the midwife who catches the baby,” thereby learning from the different people and organizations who work with the midwife. Some cautioned, however, that Black community midwives alone cannot overcome macro-level forces that are driving the Black maternal health crisis.

### Observations

#### The structure and orientation of the visits were similar

The ten-day and six-week postpartum visits began with an exchange of pleasantries, and then the midwife asked an open-ended question about what was going on with the “mama”. The midwife then conducted a detailed review of the client’s and baby’s physical wellbeing, and then the mother’s emotional, mental, and social wellbeing. The midwife reviewed both clients’ and babies’ bodily functions, nutrition, and sleep habits. There were in-depth conversations about the social support which the clients had received, who had provided it, when it was provided, and whether it was adequate. The midwife also provided encouragement and coaching on how to make requests for support. Additionally, the midwife and client reviewed how each family member—including the father—was adjusting to and bonding with the infant. The conversation also touched on how the parents’ roles and responsibilities were changing and what, if anything, was needed to stabilize the family. There was an exchange of warmth and laughter during both visits.

#### Spirituality was evident throughout the midwife-client engagement

The midwife and both clients engaged in spiritual themes during the visit. The midwife opened and closed each home visit with a brief, private spiritual ceremony, which she conducted in her car. She explained that these practices helped prepare her to serve the client and protect her health and energy after the visit. One client indicated she was “looking into her dreams” for guidance on how to name her newborn. The other client and her mother reflected on the postpartum bath ceremony that the midwife had conducted on the client a few days after the birth. The client’s mother, who was white, reported that her daughter’s ceremony had triggered the memories of her own childbirth trauma in the hospital. She tearfully revealed that she wished she had received the level of care and attention that the midwife had provided to her daughter.

#### The goals and dynamics of the visits differed

Whereas there was a significant focus on the support during both visits, there were differences in what the support was meant to accomplish. For example, the conversation in the ten-day visit focused on the support that the client needed to maintain her two-week bed confinement, whereas the conversation in the eight-week visit focused on the support that was needed for the client to care for herself, re-engage with her personal interests, and maintain her mental health as a mother of two children under two years. Additionally, whereas the midwife conducted the ten-day visit in the client’s bedroom, she conducted the other in the client’s backyard, during a ten-minute brisk walk in the neighborhood, and in the client’s bedroom.

## Discussion

To the best of our knowledge, this is the first study to take an asset-framed, multistakeholder perspective to the critically important challenge of improving Black maternal health in the United States. Our focus on “what the Black community was doing to address the crisis” employed an implicit asset frame, in that it assumed that the community was already addressing the crisis. Asset-framing has been shown to promote positive narratives and dignity, encourage collaboration and co-creation of solutions and contribute to more sustainable and culturally appropriate intervention within research populations [26]. Study participants had an overwhelmingly positive response to that frame, encouraged us to proceed with the research, and offered to stay involved. This is significant and aligns with prior research that suggest that Black people are interested in research that is solution- and community-centered and directly relevant to their lived experience. [27]

This study revealed that community midwifery provided and led by Black women is a crucial, yet under-appreciated and –resourced strategy toward this goal. The community midwifery care included medical, socio-emotional, and spiritual dimensions, focused on the themes of rest, spirituality and support, was dynamic, oriented to the entire family, and responsive to the developmental needs of the birthing person.

Midwives are highly trained health care providers who offer a wide range of essential reproductive and sexual health care services, from birth and newborn care to Pap tests and contraceptive care. These results align with prior research that shows that when midwives are central to the provision of maternal care, birthing people are more satisfied, clinical outcomes for parents and infants improve, and costs decrease [28]. For example, research conducted at CHOICES Center for Reproductive Health in Memphis, Tennessee, community-oriented midwifery clinic, showed that Black birthing parents were 2.17 times less likely to have preterm birth, 4.76 times less likely to have a low birthweight baby, and 4.55 time less likely to deliver via Cesarean section [29].

This study’s findings are reinforced by recent research that shows that midwives can help to reduce maternal and neonatal mortality and stillbirths in low- and middle-income countries. Specifically, midwifery care could avert between 22 and 65% of maternal death depending on the intensity of the investment in midwifery care [30]. A recent review by the Commonwealth Fund concluded that midwives, incorporated fully into US maternity care systems, could reduce perinatal health disparities and help address provider workforce shortages [28]. This data has been borne out in the global context, with traditional midwives acting as bridges and trusted advisors for women who might be otherwise reluctant to engage with healthcare systems [31].

The structure and content of the observed visits align with the midwifery model of care. The midwifery model takes a holistic, relationship-centered approach to the pregnancy and birthing continuum that advocates for trust and respect between the midwife and childbearing person, prioritizes and encourages parent’s autonomy, self-determination and satisfaction, allows for partnership in informed decision-making with the childbearing person, and supports a sense of safety and assurance for the birthing person and midwife [28]. Additionally, the observed midwife-client encounter extended to 60 days beyond the 42-day postpartum milestone visit that mainstream medical providers recommend [32]. Our study highlights the importance that the midwifery model of care places on the postpartum period, a period that Black women are at highest risk for death and other complications [33].

The structure and content of the observed visits, however, may go beyond the typical midwifery model. The themes of rest, support, and spirituality during the visits as well as the laughter and warmth expressed during the visit suggest a sense of intimacy, authenticity between the birthing person and midwife. While developing new birthing safety paradigms, Karen Scott described the concept of “sacred birth” i.e., that Black birth is inherently normal, healthy, spiritual, familial and more. Put simply, healthcare should seek to “advance the power and potential, not pathology, of Black people” [34]. Our study’s findings underscore the importance of the research conducted by Karen Scott and others that highlight the importance of emotional safety in obstetric care [35].

Existing midwifery models of care advanced by midwives Jennie Joseph and Jamarah Amani, JJ Way and the 21-Point Black Midwives Care respectively, exemplify the importance of this more expansive perspective by recommending that providers offer “wraparound care” which includes informing birthing people about all their birth options, involving family members in perinatal care, and honoring the cultural beliefs or traditions the birthing person holds [36, 37]. The observed rapport between the client and midwife may be a factor that mediates the positive effects of culturally competent/congruent care and may serve as a protective factor against stress for Black birthing people during childbirth and the postpartum period. Our study also aligns with research on the importance of social support networks as a protective factor in Black childbirth. Social networks provide emotional and logistical support and help birthing people feel less isolated and more empowered during pregnancy [38].

These findings, however, also provide a critique of the harmful narratives that focus on deadly maternal health statistics without a commensurate attention to the solutions, especially the outsized positive impact that Black women are making in the maternal health landscape. Our work demonstrates that Black midwifery is focused on centering Black joy, safety, community and dignity in pregnancy. Prior reports have documented Black women’s demonstrated leadership in diverse ways: hosting community baby showers; establishing community midwifery schools, community midwifery quality improvement institutions, and midwife collaboratives; building new community-owned birth centers; establishing birth center networks; expanding longstanding home births practices into birth center practices; and working on integrating midwifery into hospital institutions [39–41].

Moreover, our study also provides a critique the lionization of Black women leaders, including midwives, by mainstream society without meaningful support and investments in them and in their work. It also bolsters the national calls by Black community leaders, most of them women, who have been advocating for investments in community-based childbirth, including community midwifery as a practical and moral solution to the current maternal mortality crisis [42]. Our study also re-affirms the importance of highlighting black joy and focusing on community assets.

Black community midwives may amplify the factors that are known to be protective by providing a comprehensive care model that addresses Black birthing people’s medical and socio-emotional and spiritual needs. Our findings support the national movement led in part by Black women to increase opportunities for community birth through the proliferation of community birth centers by and for Black and people of color communities and community midwifery schools. Birth Center Equity and the Commonsense School of Midwifery are part of that movement [43, 44].

Black communities, however, face several pressing barriers to accessing Black community midwifery. First, there is poor access to community midwives because midwifery care is not covered by insurance in most states, and those who want that care face high out of pocket costs. Black childbearing people experience the biggest gap between the demand for and access to Black community midwifery [28]. Second, Black midwives in their quest to serve their communities face funding, legislative and regulatory hurdles. These include states’ restrictions of their autonomous practice, beggarly Medicaid reimbursements, and lack of access to federal funding for education and training. This study suggests that there should be more programmatic, policy, and research investments in Black community midwives, as transformative providers, to address the Black maternal health crisis.

### Limitations

Our findings are based on conversations that sought to understand the Black community’s response to its maternal health crisis to develop a research study that would meaningfully contribute to that community’s efforts. Although this study provides several important insights, there are several limitations that need to be acknowledged. First, the snowball sampling approach limited the study to a convenience sample of those who were already connected to participants, agreed to be interviewed or had the time to talk to us. We missed the perspectives of those who were not already connected to those who had been interviewed. Second, all conversations were conducted in English and non-English speaking Black communities in the United States were not included in this study. Third, we conducted two observations; we observed one midwife with two different birthing people. The observations were remarkable in their scope and relevance to the contemporary challenges of Black childbirth and postpartum care; however, we cannot assess how commonplace this midwife’s approach to care is among Black midwives. Fourth, we did not include birthing people to understand their experiences of Black community midwifery care and we may have missed issues that are important to them.

This study lays a strong foundation for future research regarding Black community midwives’ experience providing care to Black birthing people and the impact of that care on the birthing person, their families, and the community. Additional research is also needed to better understand the experiences of Black birthing people and their motivations for engaging Black community midwives. Next steps include a fuller exploration of the care that Black midwives provide to Black birthing people, the experience of Black midwives serving Black communities, and the impact of that care on the birthing person, families and communities.

## Conclusion

Our study provides an important critique of the current approach to addressing the Black maternal health crisis and a clear recommendation that investments in Black community midwifery are an essential component to addressing it. The perspectives that we captured are absent from the empirical literature and provide important insights about community-based births among Black birthing people.

## Data Availability

We are currently approved by our IRB office to share data only in aggregate. We will work with our IRB to comply with the recommendations.

## Acknowledgments

We sincerely thank the individuals who participated in the study and spoke candidly about the challenges and opportunities that the Black maternal health crisis presents. We also are indebted to the midwife and her clients who agreed to be observed in their homes. And we are sincerely thankful to Chinmayee Balachandra for her support in developing the manuscript.

